# Acquisition and onward transmission of a SARS-CoV-2 E484K variant among household contacts of a bamlanivimab-treated patient

**DOI:** 10.1101/2021.10.02.21264415

**Authors:** Arick P. Sabin, Craig S. Richmond, Paraic A. Kenny

**Affiliations:** Department of Infectious Disease, Gundersen Health System, La Crosse, Wisconsin; Kabara Cancer Research Institute, Gundersen Medical Foundation, La Crosse, Wisconsin; University of Wisconsin School of Medicine and Public Health, Madison, WI

## Abstract

The implementation of monoclonal antibody therapeutics during the COVID19 pandemic has altered the selective pressures encountered by SARS-CoV-2, raising the possibility of selection for variants resistant to one or more monoclonal antibodies and subsequent transmission into the wider population. Early studies indicated that monoclonal antibody treatment in immunocompromised individuals could result in within-host viral evolution preferentially affecting epitopes recognized by these antibodies, although whether this signifies a real risk of transmissible antibody resistant virus is unclear.

In this study we have taken advantage of a regional SARS-CoV-2 genomic surveillance program encompassing regions in Wisconsin, Minnesota and Iowa to monitor the introduction or de novo emergence of SARS-Cov-2 lineages with clinically relevant variants. Here we describe a newly acquired E484K mutation in the SARS-CoV-2 spike protein detected within the B.1.311 lineage. Multiple individuals in two related households were infected. The timing and patterns of subsequent spread were consistent with de novo emergence of this E484K variant in the initially affected individual who had been treated with bamlanivimab monotherapy. The subsequent transmission to close contacts occurred several days after the resolution of symptoms and the end of this patient’s quarantine period. Our study suggests that the selective pressures introduced by the now widespread administration of these antibodies may warrant increased genomic surveillance to identify and mitigate spread of therapy-induced variants.

## INTRODUCTION

Emergence of SARS-CoV-2 mutations spread across multiple viral lineages capable of increased transmissibility or producing reinfection despite vaccination has caused recent concern [1, 2]. In addition, the deployment of several targeted monoclonal antibody therapies [3] and vaccines [4] has introduced numerous selective pressures not previously encountered by SARS-CoV-2. For each of these reasons, improved genomic surveillance will be critical to monitor the spread of these new variants and detect the emergence of new lineages with similarly concerning properties.

Since March 2020 we have monitored the introduction and spread of SARS-CoV-2 to the service area of the Gundersen Health System, an integrated healthcare system headquartered in La Crosse, WI and providing care in 21 counties in southwestern Wisconsin, northeastern Iowa and southeastern Minnesota. We have performed viral whole genome sequencing on more than 1,900 positive cases from this region over the span of the pandemic. Among our major goals were monitoring for introduction of viral variants into our service area that had been recognized elsewhere by genomic surveillance measures as variants of concern. In kind, our program enabled early detection and tracking of clinically relevant polymorphisms in lineages in which they had not previously been identified elsewhere.

The E484K variant in the Spike protein has emerged numerous times in different SARS-CoV-2 viral lineages, including in several emerging variants of concern: the B.1.1.7 variant first identified in the UK [5], the B.1.351 variant first identified in South Africa [6] and the P.1 variant first identified in Brazil [7]. This suggests that convergent evolution toward some transmission-favoring phenotype may be occurring. The spike protein plays a key role in viral entry [8, 9] and is also the target of both naturally developed antibodies [10] as well as synthetic monoclonal antibodies used as treatment or post-exposure prophylaxis [11]. Studies have indicated that viruses bearing E484K are associated with a diminished response to vaccine-induced neutralizing antibodies [12].

Therapeutic monoclonal antibodies received Emergency Use Authorization (EUA) in the outpatient setting following trials demonstrating a modest reduction in the risk of hospitalization [13-15]. Initial authorization of these agents accounted for a drop in clinical utility if the prevailing circulating viral substrains shifted to include variant(s) for which these antibodies had reduced affinity, with continued availability to be based on passive monitoring of sequence data accumulating in repositories. Epidemiologic trends subsequently lead to the withdrawal of Bamlanivimab [16] (and later Bamlanivimab/Etesevimab [17]) from widespread clinical use, though the combination was later reinstated for widespread use and expanded for use as post-exposure prophylaxis as the prevailing circulating variants changed once again [18]. Casirivimab/imdevimab became available under EUA on 11/20/20 [19] and use for post-exposure prophylaxis was subsequently authorized [20].

The potential for these treatments to select for emergence of antibody resistance mutations has been noted previously in closely monitored immunocompromised patients [21-23], but post-administration surveillance in either close contacts or the broader community has not been reported. Between Nov 2020 and September 2021, our program administered monoclonal antibody therapies of Bamlanivimab, Bamlanivimab/Etesevimab, or Casirivimab/Imdevimab to 1,043 COVID-19 positive patients. In parallel, our regional SARS-CoV-2 sequencing program [24-26] provided the opportunity to detect newly emerging E484K-containing lineages and ascertain the potential epidemiological association, if any, with individuals receiving monoclonal antibody therapy.

## MATERIALS AND METHODS

### SARS-CoV-2 sequencing and analysis

cDNA was generated from residual RNA from diagnostic specimens using ProtoScript II (New England Biolabs, Ipswich, MA). The Ion AmpliSeq SARS-CoV-2 Panel (Thermo-Fisher, Waltham, MA) was used to amplify 237 viral specific targets encompassing the complete viral genome. Libraries were sequenced and analyzed as we have previously described [24-26]. For phylogenetic inference (i.e. to determine the hierarchy of case relationships), sequences were integrated with associated metadata and aligned on a local implementation of NextStrain using augur and displayed via a web browser using auspice.

### E484K mutation rapid tests

#### A. PCR/Restriction Digest test

cDNA from SARS-CoV-2 specimens was amplified using the following primers which span the G23012A polymorphism which encodes E484K: CTTGATTCTAAGGTTGGTGGT and GTAAAGGAAAGTAACAAGTAAAACC. The resultant 157 base pair products were digested with the MseI restriction enzyme which cuts the variant but not the reference allele. Genotypes were determined by band size discrimination using agarose gel electrophoresis.

#### B. TaqMan Assay

A custom TaqMan SNP genotyping assay (ThermoFisher) was designed to discriminate G v A alleles at 23012. Primers used were GCCGGTAGCACACCTTGT (forward) and GGGTTGGAAACCATATGATTGTAAAGG (reverse). Reporters were AATGGTGTTGAAGGTTT (VIC, reference allele) and AATGGTGTTAAAGGTTT (FAM, variant allele). The assay was run on a Biorad IQ5 real-time PCR machine and NGS-verified reference and variant samples were used to validate allele discrimination.

### Ethical approval

Specimens were analyzed in this study under a protocol approved by the Gundersen Health System Institutional Review Board (#2-20-03-008; PI: Kenny) to perform next-generation sequencing on remnant specimens after completion of diagnostic testing.

## RESULTS

While performing sequence surveillance for SARS-CoV-2 variants of potential clinical significance, we sequenced a case from Houston County, Minnesota sampled in late December 2020 which contained a genomic variant, G23012A, encoding an E484K mutation in the spike protein. Lineage analysis using Pangolin [27] assigned it to B.1.311, a lineage in which Spike:E484K had not been previously detected. This genome was from a child, the third person diagnosed with COVID-19 in a household, whom we designate “P3”. Given the then recently emerging concerns surrounding multiple other lineages containing this variant [6, 28], we analyzed household contacts of this case.

The first case in this family, “P1,” developed symptoms of COVID19 on a date in the first half of December which we designate as “Day 0” and number other events accordingly. P1 was diagnosed with a rapid COVID19 test on Day 1. Because this was a rapid test, no residual sample was available for the sequencing team. P1 was treated with infusion of bamlanivimab on Day 2. Their disease course was unremarkable and quarantine was discontinued on Day 10. Notably, P1’s electronic medical record provided no indication of any known immunocompromising condition.

P1’s spouse (P2) tested negative on Days 1, 16 and 18 but developed symptoms and tested positive on Day 23, 13 days after P1 discontinued quarantine. There were two children in the household, designated P3 and P4. P3 developed symptoms of COVID19 (day 24) and tested positive (Day 25). The second child (P4) was also symptomatic on Day 24, tested negative on Day 25 but was positive upon repeat testing five days later. No residual specimen was available from P2 or P4, but the sample from P3 was sequenced as part of our regional surveillance, and was found to have the E484K-encoding mutation. Two members of the extended family, living together nearby, also developed symptoms. P5 and P6 developed symptoms on Days 26 and 27, respectively. Both tested positive on Day 30 and received bamlanivimab infusions on Day 32. Sequencing confirmed that both P5 and P6 had the E484K variant. A summary of the timeline is shown in Figure 1.

**Figure 1.**
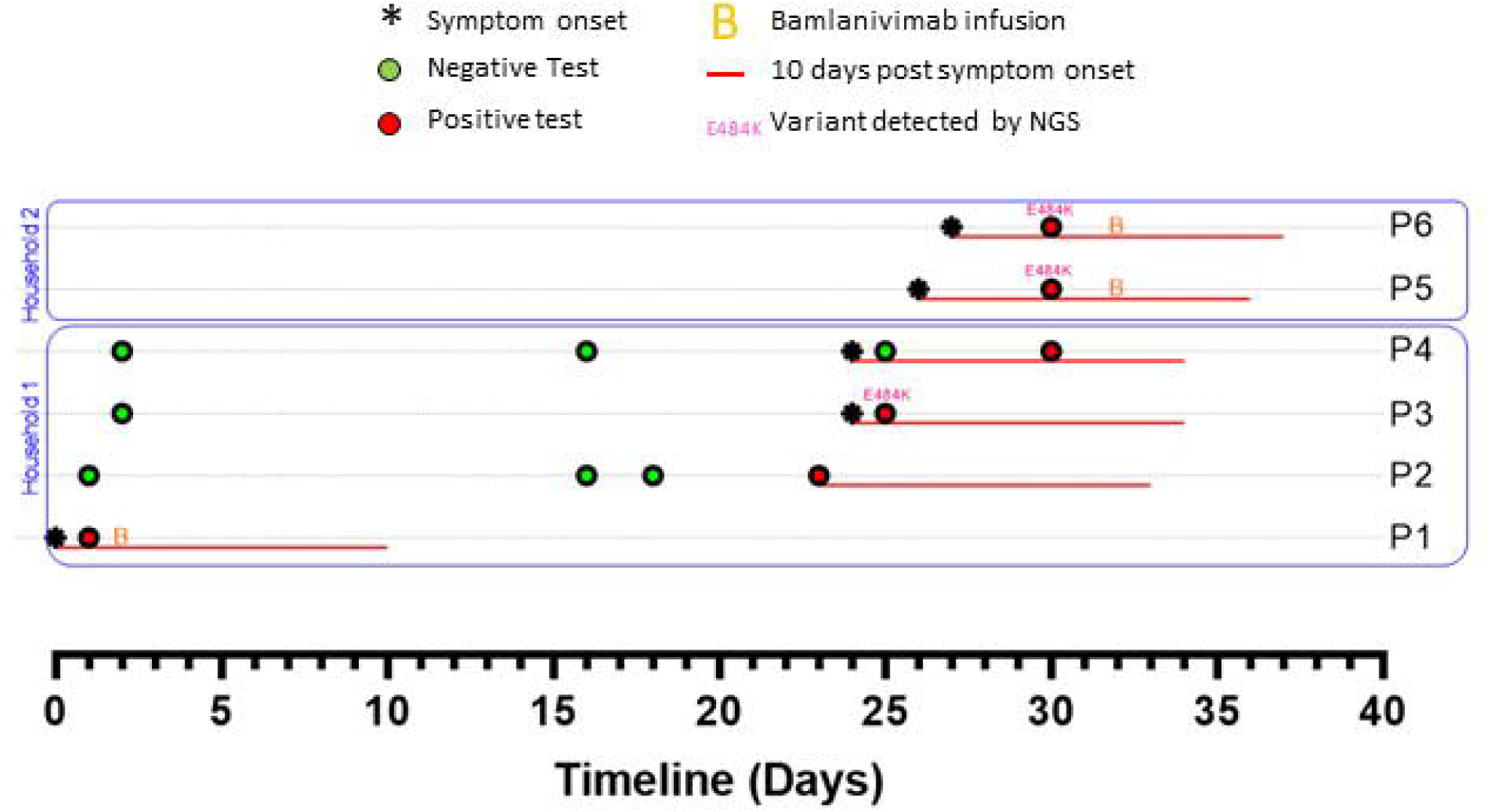
Timelines of SARS-Cov-2 testing, symptoms and treatments among household and close contacts of a bamlanivimab-treated patient (P1). Genome sequences for P3, P5 and P6 correspond to GISAID accession IDs EPI_ISL_830649, EPI_ISL_861473 and EPI_ISL_861472.

The observed transmission pattern was consistent with the possibility that the E484K variant emerged in P1 following bamlanivimab treatment, however unambiguously demonstrating this was not possible as we lacked a specimen from this case for sequencing. Nevertheless, several lines of evidence were consistent with this possibility:

Firstly, despite widespread regional surveillance, we did not detect this B.1.311/E484K lineage in any patient except those associated with this particular household.

Secondly, we sequenced a case from an adjacent county (WI-GMF-48798) which was the immediate viral ancestor on the B.1.311 lineage to this new E484K containing strain (i.e. it was identical to the three sequenced genomes with the exception of the G23012A mutation encoding E484K and the subsequently acquired variants that distinguished the three household contacts (Fig 2). Thus, while we cannot formally exclude the possibility that the E484K substrain and its immediate ancestor BOTH emerged elsewhere and arrived in our region in parallel, it seemed plausible that the E484K containing variant of this lineage may have originated locally.

**Figure 2.**
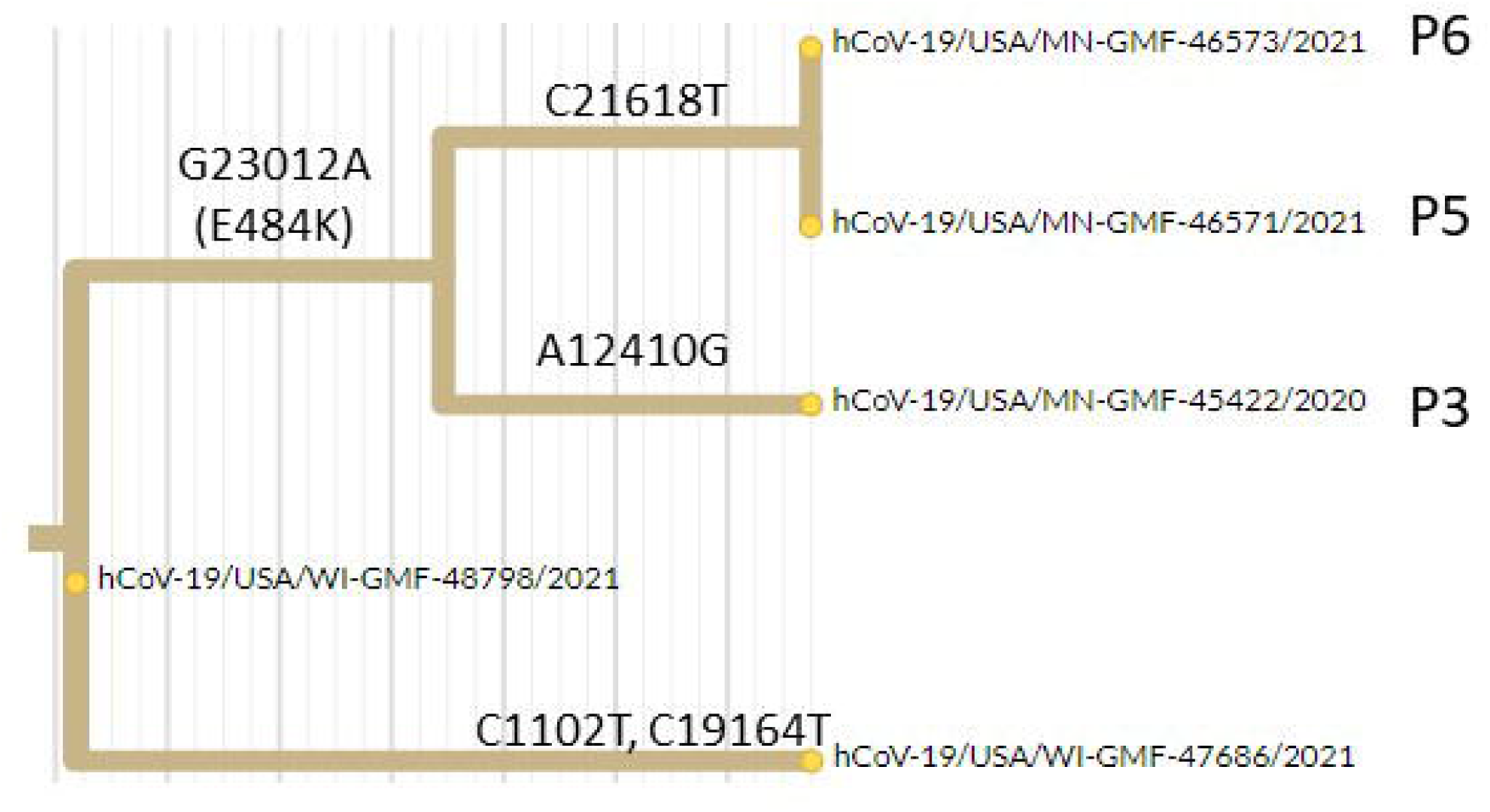
Hierarchical relationship between sequenced B.1.311 genomes related to this transmission chain. Excerpt from phylogenetic tree showing acquisition of the E484K encoding mutation (G23012A) on the branch leading to this cluster (P3, P5, P6). A local ancestral sequence to this cluster from an adjacent county is hCoV-19-USA/WI-GMF-48798/2021 (EPI_ISL_942822) and an independent lineage arising from that ancestral strain in the same county is hCoV-19-USA/WI-GMF-47686 (EPI_ISL_942808).

Third, the time lag between the presumed resolution of P1’s infectious period and the onset of symptoms (denoting an active infection) in other household members was consistent with a span of ongoing viral replication under selective pressure from the enduring monoclonal product remaining in the source patient’s circulation. During this period, presence of persistent meaningful antibody titers in the wake of ongoing viral replication may have been sufficient to incite selection of fit subpopulations of virus harboring de novo E484K and other mutations from which transmission to new hosts in close personal contact subsequently occurred.

Fourth, the close timing of symptom onset between P2-P6 makes it less likely that any of these individuals were infected for long enough to transmit the virus to one another, leaving the most likely proposition that P1 acted as the source for each of these transmission events. The number of new variants detected among the three sequenced individuals was striking for the degree of genetic diversity demonstrated by sequencing despite such a presumably short transmission chain. This was notably more diversification than we observed in multiple studies of comparable outbreaks of a similar time scale in other households, workplaces, or congregate settings such as skilled nursing facilities (data not shown). The P5 and P6 viral genomes were identical to each other but distinct from P3, ruling out a direct transmission in either direction between P3 and P5/P6. Because P5 and P6 symptom onset dates were only one day apart, it seems likely that they were infected by a common source rather than transmitting one to the other. The most likely explanation is that each acquired infection from a nearby source with considerable internal viral evolution.

Having detected this potential concern in our institutional monoclonal antibody program, we quickly sought to develop the tools necessary to establish the incidence (de novo E484k mutations) and prevalence (propagation of these mutations within a chain of infections in close contacts) of this phenomenon in hope that we would have the ability to quickly test either patients (for treatment selection) or close contacts (for resistance variant detection). NGS testing does not have the throughput or turnaround time to be useful for this purpose. Two rapid assays were developed, validated in specimens of known genotype, and are described in the Methods. One method relies on differential restriction digest sensitivity of the mutant allele (making it suitable for lower resource settings and for potentially screening for variants in pooled specimens) and the other was a Taqman assay permitting differentiation of the E and K alleles at this locus (Fig 3).

**Figure 3.**
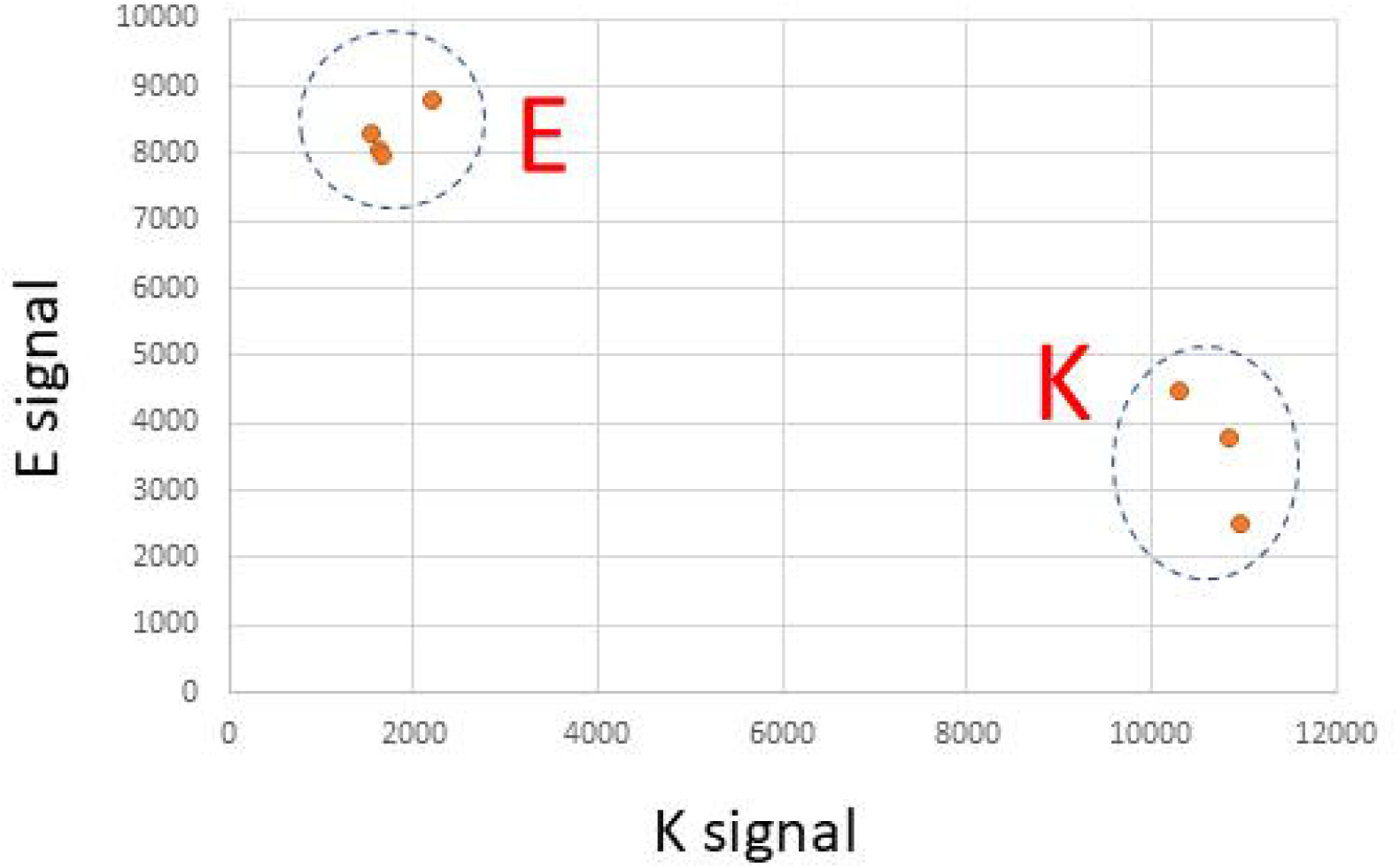
Validation of rapid Taqman SNP genotyping assay for the G23012A polymorphism. Clear discrimination between the G allele (encoding E at Spike:484) and the A allele (encoding K at Spike:484) is shown.

To evaluate this testing strategy, we reviewed all patients diagnosed with COVID-19 at our central laboratory between 12/8/2020 and 1/14/2021 and determined which of those belonged to households in which a COVID-19 positive individual had already been treated with bamlanivimab. Of 257 monoclonal antibody-treated patients, we identified 77 which had additional positive cases at the same address. Of these, 5 cases were diagnosed after the bamlanivimab-treatment date (range 1 -7 days). Using our rapid tests, we genotyped the household contact specimens. 2/5 specimens yielded no detectable signal (these had average CT values of 33 and 37 on the diagnostic test), while the remaining three specimens were clearly wild-type at this position. Notably, most of these cases demonstrated a narrow window of opportunity for infection between the two household diagnosis dates (range 1 – 4 days) and thus were particularly unlikely to have succumbed to infection with a virus that evolved under a period of prolonged mutagenic pressure in the mAb-treated host. Nevertheless, we believe that once particular nucleotide variants of concern are established, the more widespread deployment of rapid assays such as this one will offer significant benefits in early detection and classification compared to the slower speed and greater infrastructural, logistical, and financial requirements of NGS.

Although broad, our regional surveillance did not have complete coverage of our population leaving open the question of whether additional onward transmission from this cluster might have occurred or whether viral genomes sequenced elsewhere might support or refute our hypothesis. Allowing time for more genomic data to accumulate and be submitted to repositories, we revisited this question in August 2021. We obtained the 3,061 genomes from the B.1.311 lineage that had been deposited in GISAID at that time and plotted their relationships on a radial phylogenetic tree. We identified four independent emergences of variants at spike E484. In addition to the E484K described in our Minnesota cluster, we found examples in New York (1 sequenced case) and California (2 cases), and an E484Q case in Maryland. No additional genomes on the Minnesota E484K sub-lineage were detected in this analysis or our own longitudinal surveillance. Accordingly, while we cannot exclude any further transmission, it seems unlikely that this particular sub-lineage went on to seed a very large outbreak elsewhere in our region.

## DISCUSSION

In this study, we describe the probable emergence of a de novo E484K mutation and subsequent transmission to all members of a household and then onward to additional close interpersonal contacts. While the potential for this to occur has been clear for some time in immunocompromised patients, we believe that this is the first genomically-supported demonstration of subsequent transmission of a de novo mutation of concern following the probable selection for this mutation after monoclonal antibody therapy administration to the index case.

Widespread availability under emergency use authorization (EUA) has lowered the barrier to use of monoclonal antibodies, however this has generally not been paralleled by expanded utilization of sequencing approaches to monitor the impact of these treatments on the circulating repertoire of variants. As almost all public health sequencing is performed on deidentified specimens, data on viral strains and epidemiologic connections to specific individuals with prior monoclonal antibody exposure are challenging to link. Our center has administered over one thousand doses of monoclonal antibody products and was contemporaneously an early adopter of SARS-CoV-2 genome sequencing, aided in part by an IRB waiver allowing review of patient records for parameters relevant to COVID-19 disease epidemiology across our service area. This enabled an unusually high-fidelity view of both local transmission dynamics as well as evolving genome-level trends throughout the course of the pandemic. Importantly, it offered the opportunity to identify clusters of linked cases, such that cases arising subsequent to an intervention (e.g. monoclonal antibody treatment, vaccine administration, amended infection control practices, etc.) were straightforward to identify.

It is unclear to what degree the issuance of the original EUA was balanced against the estimation of the hazards of inducing or propagating resistant variants in the community. Early trials suggested NGS-detectable resistance mutations in specimens from as many as 10% of treated patients [13], although the potential for onward transmission was not explored. Indeed, detection of such mutations in an individual with a sensitive NGS assay does not necessarily imply that replication/infection-competent virus is being produced at levels sufficient to induce a clonal infection of an exposed close contact. In our study, the transmission pattern observed in these households suggests that that is precisely what has occurred.

Though single-agent monoclonal antibodies were introduced into widespread use first, the majority of the products being administered are combination products involving mixtures of antibodies that should, in theory, reduce selection for mutations of concern. In principle, this should concomitantly reduce the overall probability of randomly selecting a resistance variant with de novo in cis mutations at two distinct epitopes. Lastly, it would provide therapeutic durability for the agents by maintaining at least partial efficacy in the event that viral epidemiology shifted to include dominating variants with a single mutation at one of the two designated epitopes.

However, caution is warranted for the latter situation, as virus already resistant to one of the two cocktail antibodies would reduce barriers to resistance for emergence of a variant that may demonstrate resistance to both antibodies. Pre-testing patients for resistance-determining SNPs using rapid assays prior to antibody treatment would allow for clear contraindication if resistance to both antibodies is already present as well as permit a more nuanced risk assessment if the patient is carrying a strain already resistant to one antibody in the cocktail. Although mixing and matching antibodies to select an optimal cocktail from the approved agents is not permitted at present under the respective EUAs, this might in principle allow for more effective treatments for patients carrying virus of known genotype meanwhile significantly reducing the probability of emergence of resistance variants. As a second layer of protection, rapid genotyping of virus from household contacts of monoclonal antibody treated patients might allow more rigorous isolation methods to be used for those found to be carrying resistance variants, and therefore prevent their spread outward into the broader community At present, no clear mechanism exists for detecting small pockets of meaningful resistance in this manner unless coincidentally captured by the surveillance tools of a sentinel laboratory.

Widespread use of monoclonal antibodies, especially when regarded as a preference by individuals over vaccination, is suboptimal for several reasons: monoclonal antibodies do not prevent transmission in the very near term, do not cure the recipients immediately, provide no clearly durable immunity against reinfection, are linear interventions that scale poorly to populations experiencing exponential disease transmission, and require processes for administration that may divert limited resources from other critical healthcare infrastructure during a crisis. Our study adds the concern that such widespread use may additionally lead to the selection of antibody resistant strains which might also have some cross-resistance against prior natural or vaccinal-acquired immunity. Substantially improving surveillance in the context of monoclonal antibody utilization would give a better assessment of the likelihood of transmission events such as we describe, though, even if appropriately quantified, the risks might not be outweighed by the potential benefits of the antibody intervention itself.

In the absence of a predictable rate by which such mutations occur following monoclonal antibody administration, it is unclear what thresholds would need to exist to guide decisions regarding appropriate use in a population that does not (or will not) possess the capability to conduct adequate surveillance for these mutations. Monoclonal antibodies represent a fixed epitope intervention, used in the context of a rapidly evolving and genetically vibrant landscape. NGS currently lacks the turnaround time and scalability to be routinely useful in such a dynamic environment. Development of rapid means to characterize all resistance mutations of consequence in the viruses in circulation would permit a scalable capacity to both prospectively inform the optimal use of antibodies, or in turn to discourage their use in pockets of propagation within which these interventions may compound the risk of genomic drift by way of untoward selective pressures.

In assessing these competing risks/benefits we should also consider whether and to what extent to which this particular risk influences the broader epidemiology of the pandemic in the face of unrestrained global spread. The potential for newly emerging variants such as Alpha and Delta to rapidly overtake the local strains in circulation and quickly supplant any locally-grown genetic diversity has been repeatedly demonstrated, both in our own region and abroad. Given the widespread use of these agents, if this was a very common phenomenon overall than we might expect to see more frequent examples of independent E484 mutations at clade level than are observed (e.g. Fig 4). Nevertheless, this potential should be added to the risk/benefit analysis of deployment of these treatments at large scale in the general population.

**Figure 4.**
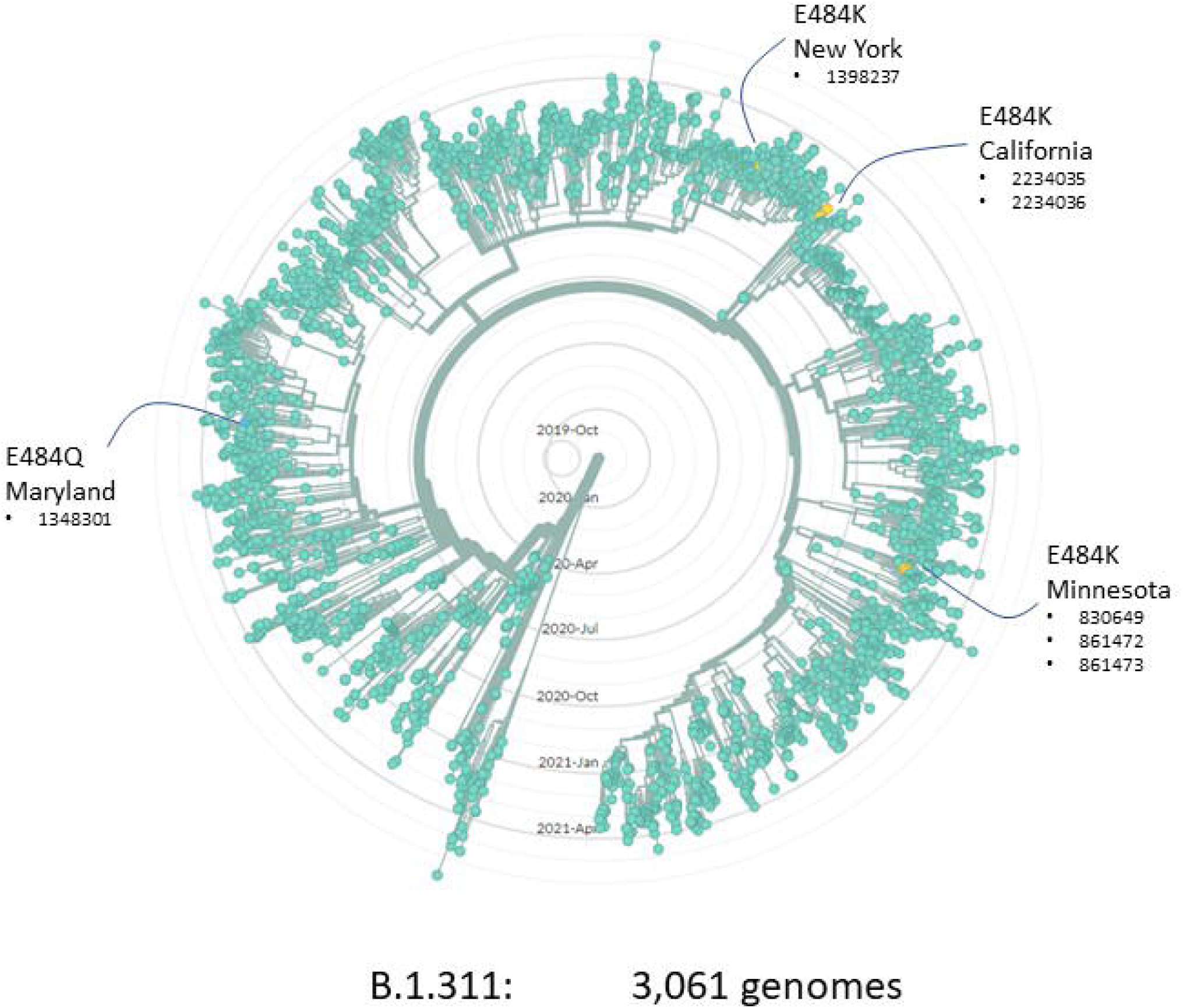
Radial phylogenetic tree of 3,061 B.1.311 viral genomes from GISAID (August 2021) showing four independent emergences of sub-lineages with mutations affecting E484. Numbers indicate the GISAID accession number (EPI_ISL_xxxxxx) for each genome.

Yet even if the spontaneous emergence of resistance variants do not tend to propagate far from their source (as in the 4 emergences on the B.1.311 lineage in Fig 3), they still have the potential to alter treatment outcomes locally, for example if resistance were to emerge in the early stages of a skilled nursing facility outbreak. In our own region, genome-level data would have suggested a continued efficacy of Bamlanivimab/Etesevimab despite a national withdrawal of its availability based on broader trends; this in turn reduced local access to a meaningful intervention among the unvaccinated population, the effect of which is difficult to quantify.

In summary, our study demonstrates the potential for onward transmission of de novo antibody resistant variants that can emerge in patients treated in the community with single agent monoclonal antibodies. Since these agents remain in widespread use as a tool to combat the pandemic, the potential for exacerbating the existing genetic diversity of SARS-CoV-2 should be accounted for in their use. Our work would support the need for additional rapid, scalable surveillance for mutations of concern to be adopted alongside the use of these monoclonal antibodies if their scale of use is to be further expanded.

## Data Availability

All sequence data are deposited on GISAID under the accession numbers listed in the manuscript.

## ACKNOWLEDGEMENTS

This work was supported by an Emergent Ventures Fast Grant (#2243) to PK and by the Gundersen Medical Foundation. PK holds the Dr. Jon & Betty Kabara Endowed Chair in Precision Oncology.

